# Characterizing Features of the Genetic Architecture Underlying Autism from a multi-ancestry Perspective

**DOI:** 10.64898/2026.02.11.26346086

**Authors:** Marla Mendes, Chen Yang Xu, Worrawat Engchuan, Brett Trost, Xiaopu Zhou, Nelson Bautista Salazar, Jack Iglar, Bhooma Thiruvahindrapuram, Liam Wallich, Thales Henrique de Paiva, Eduardo Tarazona-Santos, Bridget Fernandez, Victor Borda, Stephen W. Scherer

## Abstract

Autism spectrum disorder (ASD; MIM 209850) is reported to vary globally from 0.01% in East Asian populations to 4.36% in certain Australian cohorts. Despite high heritability estimates (61–94%), the genetic architecture underlying ASD susceptibility remains poorly characterized across diverse populations, as most genomic studies have initially focused on individuals of European ancestry. To investigate ancestry-specific genetic contributions to ASD, we analyzed whole-genome sequencing data from three independent ASD cohorts. We identified admixed ASD probands (n = 1 033) and ancestry-matched controls (n = 1 033) and performed admixture mapping (AM). AM using five continental reference populations (European, African, East Asian, South Asian, and Native American) identified five ancestry-specific ASD-susceptibility loci, including one African-related locus at 1p21.2 near *S1PR1* and four Native American-associated loci at chromosome 11q13.4. Three of these latter loci were contiguous and encompassed genes previously implicated in ASD, notably *SHANK2* and *DHCR7*, with fine-mapping identifying a significantly associated variant between the two genes (rs77695321; P = 1.52 × 10⁻⁷). The fourth Native American-associated signal at 11q13.4 overlapped the folate receptor genes *FOLR1* and *FOLR3*, with fine-mapping identifying a genome-wide significant variant (rs7950807; P = 5.21 × 10⁻⁸). A secondary admixture mapping analysis restricted to Latin American individuals, incorporating 6 487 Brazilian controls, identified 16 additional ancestry-specific loci across seven genomic regions.

## Introduction

Autism Spectrum Disorder (ASD), or autism, is a neurodevelopmental condition exhibiting extensive phenotypic heterogeneity, including varying degrees of impairment in social communication, restricted interests, and repetitive sensory-motor behaviors^1^. While some individuals with ASD develop speech, literacy, and independence, many others require high levels of support to improve educational, health, and social outcomes ^2,3^. The impacts of ASD can be exacerbated by negative life experiences, underscoring a need for early identification. ASD represents a behavioral diagnosis and involves a comprehensive assessment by specialists using observation, parent interviews, and standardized tools now with criteria from the DSM-5 (Diagnostic and Statistical Manual of Mental Disorders, Fifth Edition) to make a diagnosis. Earlier identification of ASD may allow for the implementation of behavioral interventions during key periods of neurodevelopment, which can been shown to improve long-term cognitive, social, and adaptive functioning ^2,4,5^.

Recent reports have highlighted substantial variation in ASD prevalence across global populations, with epidemiological studies reporting rates ranging from 0.11^6^-3.2%^7^ in the Americas, 0.24^6^-3.89^8^% in Europe, 0.07-1.4% in Southeast Asia^6^, and 1.2-2.9% in African populations^6^ (Supplementary Table 1). Such variability may be explained by differences in diagnostic criteria, healthcare infrastructure, reporting practices, sociocultural factors including stigma surrounding neurodevelopmental disorders, as well as geographic, ethnic, and socioeconomic conditions ^6^. However, given the high heritability of ASD, with often-cited values of 64%-91% from twin studies^9–11^, genetic risk factors are also likely contributing to prevalence differences. For example, a study in Amazonian Amerindians identified 16 variants in three ASD-associated genes (*SCN2A*, *FOXP1*, *SYNGAP1*) showing potential frequency differences compared with other populations. These investigations comprised both common variants (with allele frequencies >5% in at least one population) and rarer variants (allele frequencies <1%) ^12^. Additionally, Indigenous Australians with ASD are reported to be twice as likely to have a profound form of disability ^13^, and some ASD-related copy-number variants (CNVs) were found to be specific to Japanese populations^14^. Studies of consanguineous or founder populations cohorts with ASD are enriched for several ultra-rare novel recessive loci^15,16^.

Despite significant progress in delineating the genomic factors underlying ASD, the contribution of ancestry to genetic susceptibility remains underexplored. Analysis of data from ASD genome-wide association studies (GWASs) has focused on individuals of European descent^17,18^. While many studies included samples from participants of other ancestries, the small size limited statistical power. This bias not only limits the ability to discover variants associated with ASD, but may also contribute to downstream health disparities in non-European ancestry populations^19^. Specifically, important variants may not be detected if they are rare or absent in European populations. In addition, differences in linkage disequilibrium structure across ancestral groups can affect both fine-mapping resolution and the transferability of association signals. Other population-specific biological features, such as, haplotype structure, gene–environment interactions, and the presence of ancestry-enriched functional variants, may further influence genetic risk architecture and variant detectability across populations ^20–22^.

Trans-ethnic analyses often rely on non-European cohorts with relatively homogeneous genetic backgrounds ^23^. However, such cohorts may be limited in sample size and may not capture the full spectrum of genetic diversity relevant to complex traits such as ASD. Here, we adopt a complementary approach by applying Admixture Mapping (AM) to individuals with recently mixed genetic ancestry (Figure 1). This strategy tests whether specific genomic regions inherited from different ancestral populations, also referred to as admixture tracts ^24^, occur more or less frequently in individuals with ASD compared with controls, leveraging the mosaic ancestry structure present within each individual’s genome ^25^. By exploiting ancestry differences along the genome rather than comparing between population groups, AM has the potential to increase power to detect ancestry-specific ASD-risk locus ^26–28^. This approach is particularly advantageous in ASD, where some large non-admixed non-European cohorts exist ^16,29^, but coverage across global ancestries remains incomplete.

**Figure 1.**
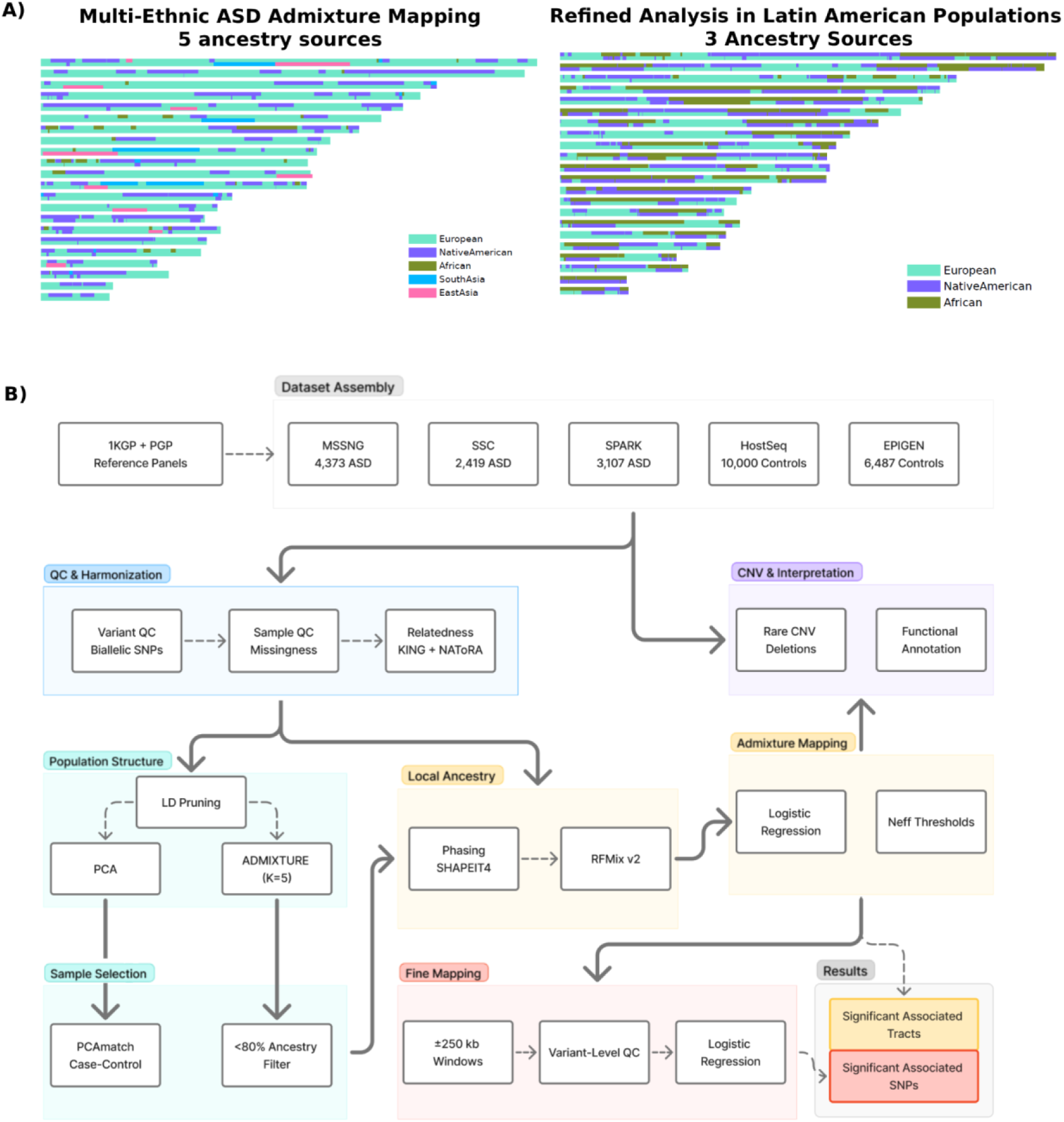
Overview of the multi-ethnic admixture mapping and integrative CNV analysis pipeline for ASD. **A)** Example of local ancestry mosaics in admixed ASD individuals. The left panel shows ancestry composition using five continental ancestry sources (European, African, Native American, South Asian, and East Asian), while the right panel shows the refined analysis restricted to three predominant ancestry sources (European, African, and Native American). Each horizontal bar represents the autosomal chromosomes of an individual, with colored segments indicating local ancestry assignments along the genome. **B)** Schematic overview of the analytical workflow used for dataset assembly, quality control, ancestry inference, admixture mapping, fine mapping, and integration of rare CNV deletions to prioritize ASD-associated loci.

We performed AM on 1 033 ASD-affected individuals with recent five-way mixed ancestry (African, European, Native American, East Asian, and South Asian) drawn from three independent whole-genome sequencing (WGS) cohorts: the Autism Speaks MSSNG Project ^30,31^, the Simons Simplex Collection (SSC)^32^, and the Simons Foundation Powering Autism Research (SPARK) cohort^33^. These individuals were analyzed together with 1 033 ancestry-matched controls from the cross-Canada Host Genome Sequencing Initiative (HostSeq) ^34^. Collectively, these cohorts largely represent North American populations, which are characterized by substantial recent admixture^35^ (Figure 2, Table 2).

**Figure 2.**
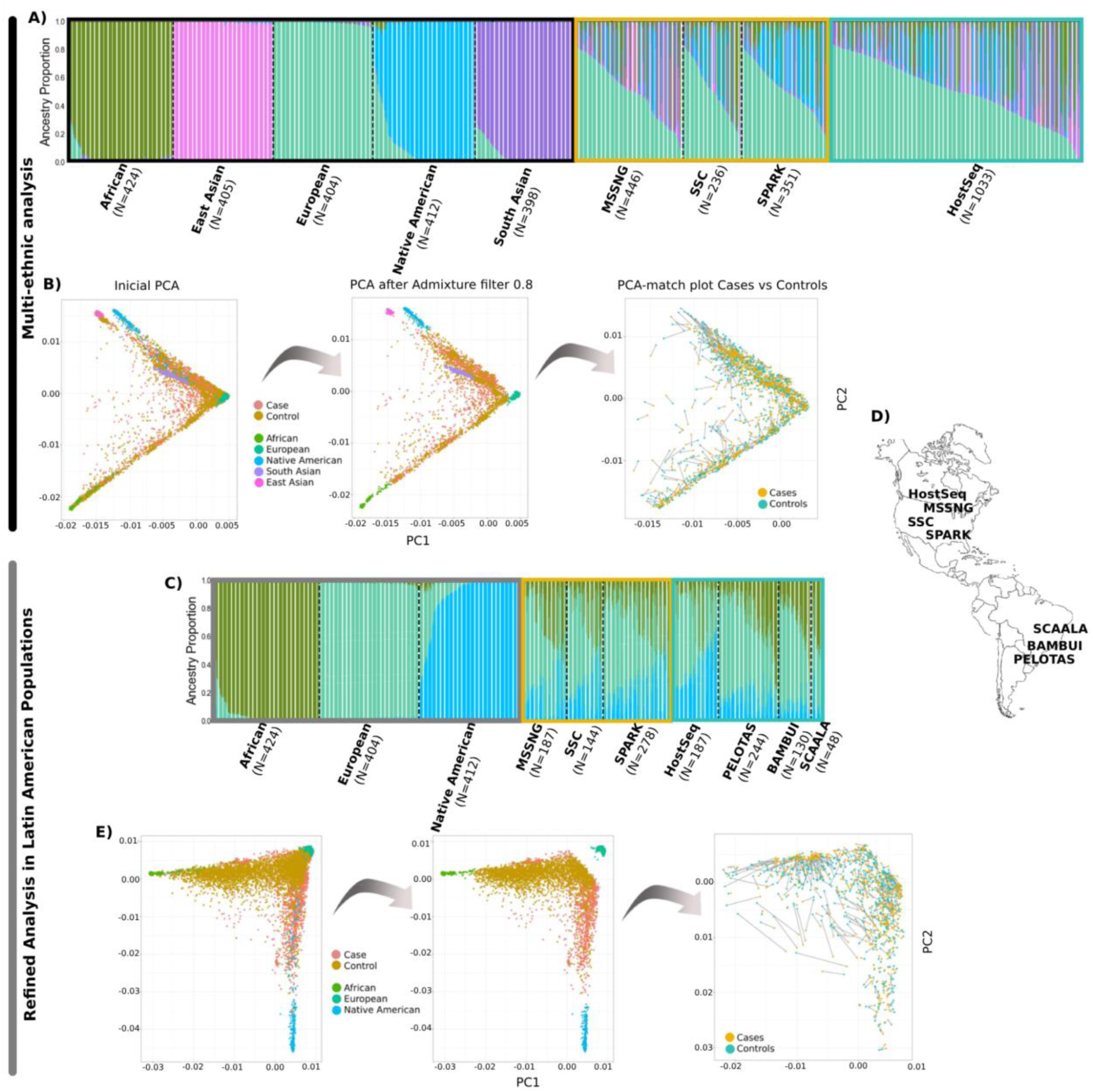
Clustering and population structure inferences. **A)** ADMIXTURE proportions per sample for the Multi-ethnic AM using five ancestral sources: African, East Asian, European, Native American, and South Asian. Each vertical bar represents the ancestry proportions for individual samples. **B)** Principal Component Analysis (PCA). The first plot shows the initial distribution of all samples. The second plot represents samples after filtering, excluding those with greater than 80% ancestry proportion from any single source based on ADMIXTURE results. The third plot, PCAmatch, demonstrates the selection of controls that most closely match the case samples based on principal component analysis. **C)** ADMIXTURE proportions per sample for the Refined Analysis in Latin American Populations, using three ancestral sources: African, European, and Native American. **D)** PCA for the Refined Analysis in Latin American Populations, following the same methodology as panel B but with a reduced set of ancestral sources (African, European, and Native American). **D)** Geographic location of each dataset, noting that geographic location does not always correspond to genetic ancestry.

**Table 2.** Initial global ancestry composition across ASD cohorts, control datasets, and reference populations. This table summarizes the average global ancestry proportions inferred for used datasets. For ASD cohorts (MSSNG, SSC, SPARK), ancestry proportions are reported just for affected samples. The lower section reports the number of individuals in each reference panel with the average global ancestry for each specific ancestry (percentage in parentheses).

|  |  |  |  |  | Average Global Ancestry Proportions |  |  |  |  |
| --- | --- | --- | --- | --- | --- | --- | --- | --- | --- |
|  | Dataset | Type | Affected Samples | Unaffected Samples | African | Native American | East Asian | South Asian | European |
| Cases | MSSNG | WGS | 4 373 | 5 075 | 6.03% | 2.42% | 5.30% | 6.09% | 80.14% |
|  | SSC | WGS | 2 419 | 6 786 | 4.86% | 2.93% | 3.81% | 3.96% | 84.42% |
|  | SPARK | WGS | 3 107 | 9112 | 5.20% | 5.13% | 3.48% | 3.21% | 82.95% |
| Control | HostSeq | WGS | - | 10 000 | 6.13% | 2.13% | 7.79% | 10.26% | 73.66% |
|  | SCAALA | Array | - | 1309 | 50.80% | 6.40% | - | - | 42.90% |
|  | Bambui | Array | - | 1442 | 14.70% | 6.70% | - | - | 78.50% |
|  | Pelotas | Array | - | 3736 | 15.90% | 8% | - | - | 76.10% |
|  |  |  |  |  | **Reference panel composition by predominant ancestry assignment** |  |  |  |  |
| Reference | 1000 Genome Project | WGS | - | 1296 | 425 (96.11%) | 64 (47.22%) | 405 (98.71%) | 402 (94.05%) | 404 (97.28%) |
|  | Peruvian Genome Project | Array | - | 358 | - | 358 (96.09%) | - | - | - |

Our goal was not only to identify ancestry-specific ASD-risk locus loci but also to evaluate their potential biological relevance. We complemented the AM with an analysis of rare CNVs deletion intersecting the newly discovered AM loci. This integrative strategy combines common-variant signals with rare, potentially high-impact variation to strengthen evidence for candidate ASD loci^36,37^. Rare CNVs data provide orthogonal evidence by highlighting local dosage-sensitive genes and regulatory elements that may contribute to ASD risk. To this end, we assessed rare CNVs in a larger ASD dataset not restricted by ancestry (n=10 263 ASD-affected individuals, referred to as “broader cohorts”) within the AM hits.

## Materials and Methods

### Datasets

We used data from the Autism Speaks MSSNG Project^30,31^, the Simons Simplex Collection (SSC)^32^, and the Simons Foundation Powering Autism Research (SPARK) cohort^33^ which collectively have genetic information for thousands of families with one or more individuals with ASD (Table 2). For controls, we used 10 000 samples from the cross-Canada Host Genome Sequencing Initiative (HostSeq)^34^. As reference data, we used samples from the 1 000 Genomes Project phase 3 data (1KGP)^38^ and 358 Peruvians from the Peruvian Genome Project (PGP)^39,40^, with Native American Ancestry (Table 2, Supplementary Information).

### Quality Control

In addition to the existing quality control procedures applied to the MSSNG, SSC, HostSeq, EPIGEN, 1KGP, and PGDP datasets, we implemented an additional set of filters specific to our AM analysis (Figure 1, Supplementary Information). We then merged the affected samples from the ASD datasets (MSSNG=4 373; SSC=2 419; SPARK=3 107) with the controls and reference data, resulting in a total of 1 533 979 variants and 21 980 samples. After inferring the kinship, we applied a second-degree relatedness filter resulting in a final dataset of 20 794 samples with 1 533 979 variants.

### Population Structure

For population structure analysis, we used the dataset after applying additional variant-level filters (Figure 1, Supplementary Information). After these filters, 228 904 variants remained. To estimate global ancestry, we ran ADMIXTURE^41^ with 10 replicates and K = 5, corresponding to five continental reference populations: Europeans, Africans, East Asians, South Asians, and Native Americans (Figure 2-A,B). We also performed principal component analysis (PCA) using SNPRelate^42^.

### Sample Selection

To prioritize admixed populations, we chose samples from both autistic individuals and control datasets, ensuring that none exceeded an 80% proportion in any ancestry source (Figure 1, Figure 2, Supplementary Information). Additionally, for the subsequent analysis, we selected samples from the reference populations (1KGP, and PGDP) with more than 80% representation from a single ancestry source to ensure their suitability. After this filter we had 1 747 autistic individuals; 1 606 controls and 2 043 reference samples (Figure 2-D). We then applied a case/control matching algorithm, selecting the closest control sample for each case using an adapted version of PCAmatchR^43^ (Figure 1-A and Figure 2-D,E). After this filter we had our final datasets for the AM analysis that had 1 033 autistic individuals; 1 033 controls and 2 043 reference samples (Figure 2-D).

### Admixture mapping

We phased our genome-wide genotype data (1 533 979 variants and 21 980 samples) with SHAPEIT4^44^ using the GRCh38 genetic map^45^. Local ancestry inference was performed using RFMix v2^46^ (Figure 1, Supplementary Information). For each genomic segment inferred by RFMix (ancestry tract), used as the unit of association testing, we quantified ancestry-specific copy number (0, 1, or 2) for African, European, East Asian, South Asian, and Native American ancestry. Ancestry tracts represent genomic regions inferred to originate from a single ancestral population based on local ancestry inference. We then used logistic regression to test the association between ASD status and the number of ancestry-specific tracts at each locus. To avoid the dummy variable trap and ensure model identifiability, we set European ancestry as the reference category and excluded it from the regression model ^47^. The association tests were adjusted for sex and the global ancestry proportions as fixed-effect covariates^48,49^ (Figure 1, Supplementary Information).

### Significance Threshold

Because our association tests are performed on ancestry tracts rather than individual single-nucleotide polymorphisms (SNPs), the conventional genome-wide significance threshold of 5 × 10⁻⁸ is not applicable^50^. To define appropriate significance thresholds for the AM analyses, we applied a Bonferroni correction based on the number of effective tests (N_eff). The effective number of tests was estimated from the total number of ancestry tracts (i.e., the unadjusted number of tests prior to accounting for correlation), scaled by the sample variance and normalized by the estimated spectral density at frequency zero^51^. The significance threshold was then calculated by dividing 0.05 by N_eff. This approach yielded ancestry-specific Bonferroni-corrected significance thresholds of 1.05 × 10⁻⁴ for African ancestry, 1.99 × 10⁻⁴ for Native American ancestry, 1.70 × 10⁻⁴ for South Asian ancestry, and 1.41 × 10⁻⁴ for East Asian ancestry.

### Fine Mapping

For fine mapping analyses, we utilized all unrelated WGS samples, regardless of ancestry, resulting in a combined dataset of 18 641 individuals (9 083 autistic individuals and 9 558 controls). Variants located within 250 000 base pairs upstream and downstream of the target regions identified in the AM results were extracted. We performed variant-level normalization and quality control on genomic data from four WGS cohorts: MSSNG, SSC, SPARK, and HostSeq, focusing on chromosomes 1 and 11. The cleaned datasets from all cohorts were merged, resulting in 5 731 variants remaining on chromosome 1 and 8 233 on chromosome 11. Association testing was performed using logistic regression in PLINK, adjusting for sex and the first five principal components (PCs) to account for population structure.

### Refined Analysis in Latin American Populations

Initial AM analyses across five reference continental populations identified multiple loci associated with ASD risk, including signals influenced by African and Native American ancestry components. To improve resolution of those signals, we conducted a refinement analysis focused specifically on Latin American individuals, who typically carry substantial proportions of both Native American and African ancestry, within an admixed background that also includes European contributions. For this, we selected individuals with self-reported (or genetically inferred when self-reporting was not available) ethnicity labeled as American, Admixed, Hispanic, Other, or Unknown. In addition, we incorporated control samples from the Brazilian EPIGEN initiative, which includes 6 487 individuals from three population-based cohorts representing diverse Latin American ancestries (Figure 1, Figure 2-C).

All previously described quality control procedures, ancestry inference methods, association testing, and significance threshold calculations were applied to this dataset. Specifically, EPIGEN is the only array-based cohort in our study, and after initial QC we retained 1 519 562 variants and 28 467 merged samples. Following relatedness filtering, 9 916 unrelated samples remained. Population structure analyses (PCA and ADMIXTURE (Figure 2-B,E)) were conducted using 84 079 variants after LD pruning and Hardy-Weinberg equilibrium filtering. We then selected only individuals whose ancestry proportions did not exceed 80% in any single ancestry component, resulting in a set of 1 505 autistic individuals, 3 214 controls, and 1 241 reference samples. PCAmatch^43^ was used to identify ancestry-matched subsets for AM (Figure 2-E). This resulted in a final dataset of 609 autistic individuals, 609 controls, and 1 241 reference samples for the Latin American AM analysis. The resulting Bonferroni-corrected significance thresholds were inferred as: African=8.50×10^−5^ and Native American=1.14×10^−4^. To enhance statistical power, we conducted a second fine-mapping analysis using also the imputed EPIGEN dataset. Prior to imputation, VCF files were assessed using the checkVCF.py^52^ script to identify and correct potential formatting and reference allele inconsistencies. When necessary, the bcftools +fixref plugin was applied to correct reference alleles. Imputation was performed on the TOPMed Imputation Server^53^ using TOPMed reference panel (version r3)^54^. Post-imputation QC followed the same pipeline as the WGS dataset, with the addition of filtering based on imputation quality score (R²>0.8). After filtering, the final dataset included 9 083 autistic individuals and 15 390 controls, with 5 287 variants retained on chromosome 1 and 7 380 on chromosome 11.

### Copy Number Variation (CNV) Deletion analysis

Recognizing the significant role of rare CNV deletions in ASD genetic architecture^55,56^, we analyzed deletions overlapping significant regions identified through AM in the MSSNG, SSC, and SPARK datasets. We focused on rare CNVs, defined as those with a population frequency of less than 1% in gnomAD^57^ using a 50% reciprocal overlap criterion to assess matches. CNVs were identified using an in-house pipeline^55,58^ involving two read-depth-based methods^59,60^.

### Biological Insights

To investigate the molecular and functional context of the AM results, including fine-mapped lead SNPs and overlapping CNV deletions, we examined the local regulatory and chromatin architecture of the associated loci. Cis-regulatory elements annotated by the ENCODE Project were visualized using the SCREEN database^61^. Additionally, we incorporated chromatin interaction data from the 3D Genome Browser to investigate the 3D genome architecture surrounding the locus^62^.

To assess whether any of these genes have been previously implicated in ASD, we incorporated two standardized gene-scoring systems widely used in autism genetics research: the SFARI Gene Score ^63^ and the EAGLE Score ^64^. The SFARI Gene Score is a categorical classification based on the strength of evidence linking a gene to ASD. Categories range from 3 (Suggestive Evidence) to 2 (Strong Candidate) and 1 (High Confidence), and also include the S (Syndromic) category. The EAGLE Score (Evidence-based Assessment of Gene Likelihood in Autism) is a continuous metric that quantify the strength of a gene’s association with ASD. Higher EAGLE scores indicate stronger and more consistent evidence across datasets, with scores ≥12 classified as “Definitive” evidence for ASD association.

## Results

### Multi-Ethnic ASD Admixture Mapping

In the multi-ethnic AM analysis, following QC and selection for recently admixed individuals, we analyzed data from 1 033 individuals with ASD, 1 033 controls matched for ancestry and 2 043 reference samples, the latter representing specific ancestral populations. We performed an association test based on local ancestry tracts, defined as the genomic segments assigned to the tested continental ancestries (African, European, Native American, South Asian, East Asian). We identified a significant association with African tracts on chromosome 1 (Table 3, Supplementary Figure 1, Supplementary Table 2, Supplementary Table 3). The associated 73.96 kb tract spans chr1:101,218,347-101,292,308 (hg38) (AM p-value=9.65×10⁻⁵) and overlaps the protein-coding gene *S1PR1* (Sphingosine-1-Phosphate Receptor 1), and the pseudogenes*, RP4-575N6.2* and *PPIAP7* (Table 2).

We also identified four significant tracts associated with local Native American ancestry on chromosome 11 (Table 2, Supplementary Figure 1, Supplementary Table 4). Three of these tracts were located in adjacent genomic intervals, while the fourth was located approximately 598 kb distal to this region (mapped to chr11:72,128,915–72,455,989, Table 3, Supplementary Figure 2-B). Together, these regions encompass several genes, including *FOLR1*, *FOLR2*, *CLPB*, *INPPL1*, *SHANK2*, and *DHCR7* (Figure 3; Supplementary Figure 1). Across this AM tracts, two genes were identified as having established evidence in SFARI and/or EAGLE (Table 2): *SHANK2*, with a SFARI score of 1 and an EAGLE score of 18.55, and *DHCR7*, with a SFARI score of S.

**Figure 3.**
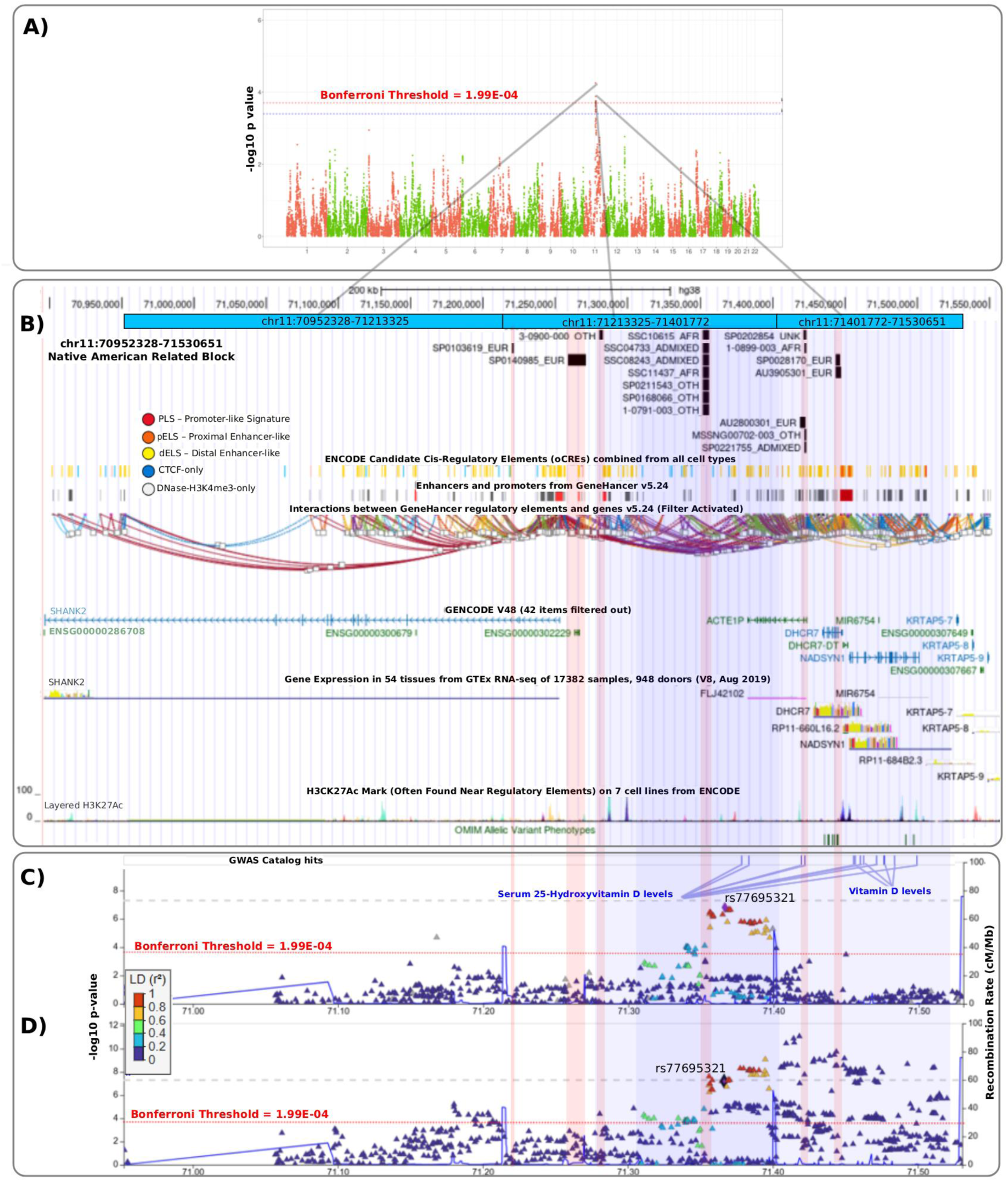
Results for Native American Ancestry for the multi-ethnic AM. **A)** Manhattan plot of AM results for 3 contiguous Native American-related ancestry tracts. **B)** UCSC Genome Browser view of CNV deletions identified in ASD probands overlapping the block (chr11:70,952,328–71,530,651). Each black bar represents a CNV deletion event, with the inferred genetic ancestry of the individual shown to the right of the sample ID: European (EUR), African (AFR), and other or admixed ancestries (ADMIXED, OTH, or UNK). Vertical red background highlights the deletion coordinates. **C)** LocusZoom plot showing the WGS-based fine-mapping results. Each point represents a SNP, colored according to linkage disequilibrium (R²) with the lead SNP rs77695321. Previously reported GWAS signals are highlighted in blue lines. **D)** LocusZoom plot showing the fine-mapping results incorporating the imputed EPIGEN dataset. The same lead SNP (rs77695321) identified in the initial fine-mapping was used as the reference for the LD color gradient. Blue backgrounds highlight the fine mapping pointed regions.

**Table 2.**
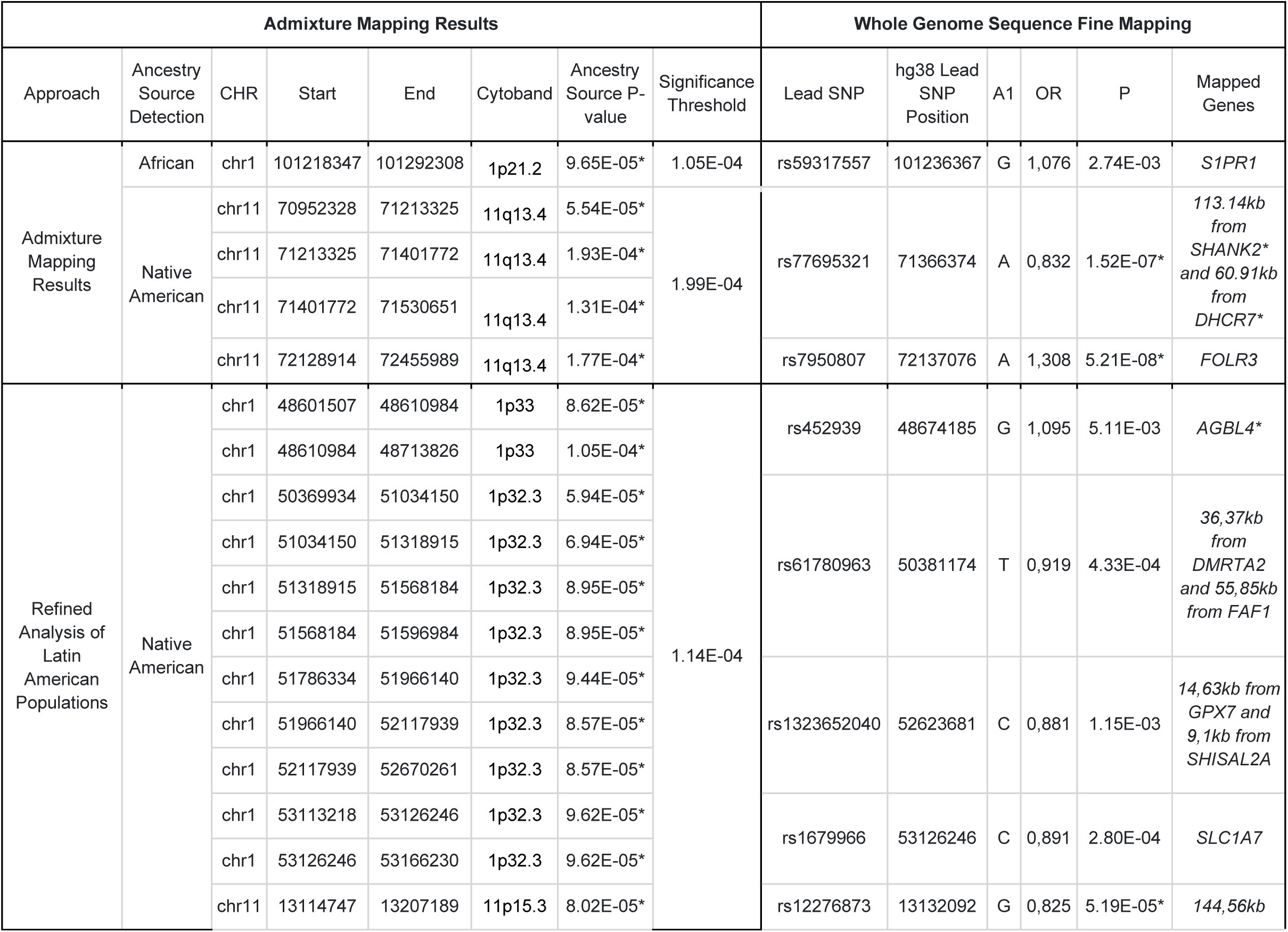
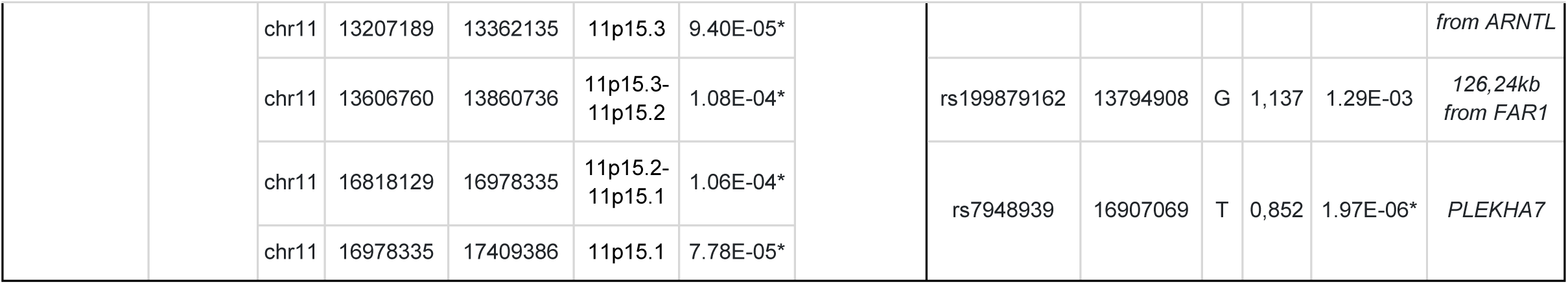
Summary of all loci identified through admixture mapping and Latin American fine-mapping analyses. P-values marked with an asterisk (*) indicate statistical significance after Bonferroni correction. Genes marked with an asterisk indicate that the gene is present in the SFARI or EAGLE dataset.

For the purposes of gene mapping and CNV overlap analyses, we grouped the three adjacent tracts into a single contiguous block of 578.3 kb spanning chr11:70,952,329-71,530,651 (Figure 3, Supplementary Figure 2-A). In this block, we detected a significant associated SNP, rs77695321 (Fine Mapping P-value =1.52×10⁻^7^), located between *SHANK2* and *DHCR7* (Figure 3-D). Within the same locus, we identified 17 CNV deletions from the broader cohorts (Figure 3, Supplementary Figure 2-A). Seven of these deletions had identical genomic coordinates (chr11:71,351,712-71,356,380 hg38 gnomAD total frequency: 5.55×10⁻⁴ ^57^, gnomAD population specific frequencies: African: 2.0×10⁻³, Admixed Americans: 7.94×10⁻⁵, 0 in all other populations ^65^). At least six of the seven are inherited (two paternal and four maternal, the remaining didn’t have inheritance information). Among the seven carriers, global ancestry estimates showed substantial heterogeneity: African ancestry ranged from 22.9% to 91.1%, European ancestry from 8.9% to 62.1%, and smaller contributions from Native American (up to 16.9%) and East Asian ancestry (up to 21.4%).

To further evaluate a potential ancestry-specific signal, we examined local ancestry inference for the ancestry tracts overlapping the deletion. Across the seven carriers, the haplotype segment overlapping the deletion was assigned to European ancestry in six individuals, while in one individual it was assigned to South Asian ancestry. Thus, despite heterogeneous global ancestry profiles, the CNV does not consistently arise on a shared ancestry tract, providing no clear evidence for an ancestry-specific haplotype origin. We also compared the frequency of this recurrent deletion across ASD cohorts and external controls (HostSeq and 1KGP). The deletion frequency was 0.06% among ASD cases, 0.04% among ASD family members, and 0.016% in external controls. However, the very small number of carriers, their diverse ancestry backgrounds, and the fact that most deletions are inherited rather than *de novo* substantially limit interpretation. As such, we cannot determine whether this event reflects an ancestry-related risk factor, a potential founder effect, or simply rare background population variation.

The fourth Native American tract was also located on chromosome 11 (chr11:72,128,914-72,455,989 hg38 p=1.77×10⁻⁴). This 327.1kb region encompasses several genes, including members of the folate receptor (*FOLR*) gene family, *INPPL1*, *PHOX2A*, and *CLPB*. Fine mapping identified the lead SNP rs7950807 (p=5.21 × 10⁻8), an intronic variant of the *FOLR3* gene (Table 2, Supplementary Table 2). In this region we identified 24 rare CNV deletions in ASD probands (1 EAS, 13 EUR, 7 OTH, 1 SAS and 2 UNK); 15 with the same coordinates (chr11:72,436,992-72,450,534, gnomAD total frequency: 4.2×10⁻⁴) (Supplementary Figure 2-B). Local ancestry inference of the 15 probands carrying the recurrent deletion showed substantial heterogeneity across continental ancestry sources.

### Refined Analysis in Latin American Populations

Given the finding in Native American and African tracts, we conducted a refined AM analysis focused specifically on Latin American individuals from the ASD-cohort, who typically carry substantial proportions of both ancestries in addition to European ancestry. This analysis incorporated 6 487 individuals from EPIGEN dataset^66^, which consists of exclusively Latin American controls. To enhance statistical power, we also performed a second Fine Mapping analysis that included the EPIGEN dataset alongside the original WGS data. This analysis identified significant SNPs in all regions previously highlighted by the initial WGS-based Fine Mapping (chr11:70952328-71530651, chr11:72128914-72455989, chr11:13114747-13362135, and chr11:16818129-17409386), and confirmed the statistical significance of the lead SNP rs77695321 (*p*=6.13×10⁻⁶; Figure 3E, Table S2). Additionally, this second Fine Mapping revealed additional significant SNPs in the AM hits on chromosomes 1 and 11 that were not identified in the initial WGS-based analysis. These discrepancies between the two analyses likely reflect differences in ancestry composition and linkage disequilibrium, particularly the higher proportion of Native American ancestry in the Latin American subset, as well as increased effective sample size and improved power when incorporating EPIGEN.

For this refined AM analysis, 16 novel significant association tracts were identified on chromosomes 1 and 11 (Table 2, Figure 4-A, Supplementary Table 2, Supplementary Figure 3, Supplementary Table 5) which were also grouped into blocks based on their consecutive genomic coordinates. The first block (chr1:48,601,508-48,713,826) overlapped *AGBL4*, a gene previously associated with autism and assigned a SFARI Gene Score of 2. Three CNV deletions overlapping this 112.3-kb block were observed in ASD probands, each occurring in individuals classified as European, African, or of unknown ancestry (Supplementary Figure 4-A).

**Figure 4.**
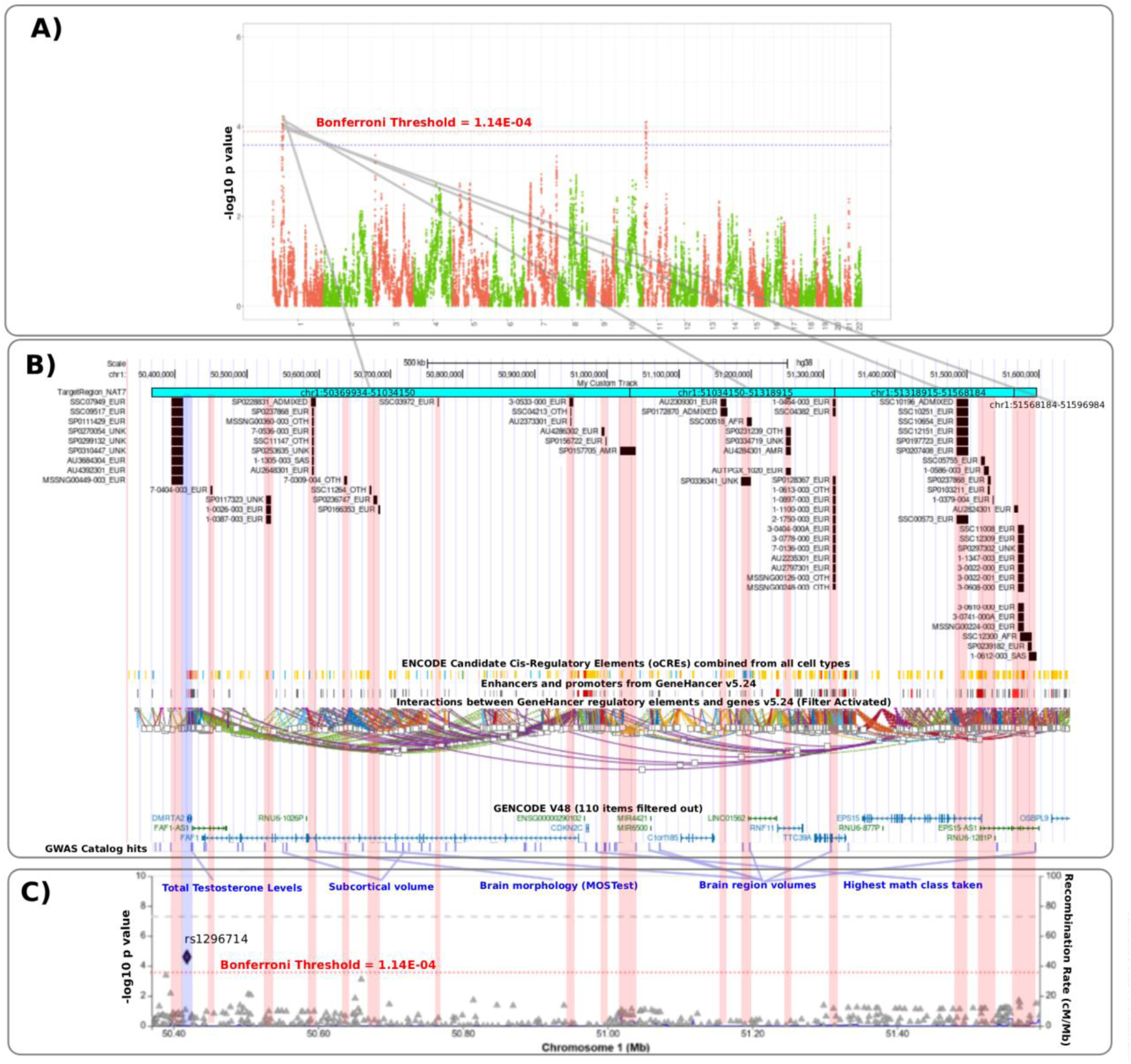
Results from the Refined Analysis in Latin American Samples. **A)** Manhattan plot showing admixture mapping results for Native American ancestry tracts. **B)** CNV deletions identified in ASD probands overlapping the region chr1:50,369,934-51,596,984. Each black bar represents a deletion event, with the inferred genetic ancestry of the individual indicated to the right of the sample ID. Vertical red background highlights the deletion coordinates. **C)** LocusZoom plot for the same region showing the fine-mapping results incorporating the imputed EPIGEN dataset. Blue backgrounds highlight the fine mapping pointed regions.

The second block (chr1:50,369,935-51,596,984) spanned several genes, including *FAF1*, *CDKN2C*, *RNF11*, *TTC39A*, *EPS15*, and *OSBPL9* (Figure 4-B). This 1.23Mb region includes multiple GWAS Catalog hits associated with brain-related phenotypes such as subcortical volume^67^, brain morphology^67^, brain region volume^68^, and highest math class taken^67^ (Figure 4-B). Notably, this block overlaps with 80 CNV deletions observed in ASD probands. In comparison, gnomAD presents 11 deletions on this same region with a frequency of 2.154×10⁻⁶ ^69^. While the presence of 80 ASD individuals with deletions in this region may add further evidence of a potential contribution of this locus to ASD risk, whether through gene dosage effects or disruption of regulatory elements, interpretation remains limited. These CNVs occur across individuals with diverse global ancestry backgrounds, and most events are inherited rather than *de novo*, reducing confidence in their pathogenic relevance.

## Discussion

Using our initial AM scan in five-way admixed WGS from individuals with ASD, we identified one African ancestry-associated tract on chromosome 1 and four Native American-associated tracts on chromosome 11. These four Native American signals clustered into two broader regions: a larger block spanning chr11:70.95-71.53 Mb, composed of three contiguous tracts, and the fourth tract located at chr11:72.13-72.46 Mb. We then refined the analysis by restricting to Latin American individuals and incorporating 6487 individuals from the EPIGEN cohort, which increased statistical power and improved the resolution of Native American ancestry tracts. This analysis detected 16 additional tracts: 11 on chromosome 1 and five on chromosome 11, which grouped into seven broader blocks based on their contiguous genomic coordinates.

In total, across both analytic stages, we identified 10 ancestry-associated genomic blocks, obtained by merging adjacent significant loci (Table 2, Supplementary Table 2). Of these 10 genomic blocks, 4 contained a significant lead SNP identified through WGS fine mapping (Table 2). In addition, 8 of the blocks overlapped rare CNV deletions found in ASD probands (Supplementary Figure 2, Supplementary Figure 4, Supplementary Figure 5). Collectively, these regions span a total of 148 genes (Supplementary Table 7).

The only gene identified in our analyses as associated with African ancestry was *S1PR1* (sphingosine-1-phosphate receptor)(p-value=9.65×10⁻⁵), which has previously been implicated in pain perception^70^. Through integrated transcriptomic analysis of ASD samples, Fan *et al.* (2024)^71^ reported *S1PR1* as being up-regulated. Interestingly, abnormality in pain perception and reactivity is often observed in people with ASD, often presenting as hyper- and hypo-sensitivity to sensory stimuli^72^. Additionally, it seems that the molecular mechanism through which *S1PR1* mediates pain insensitivity is through increased activation of KCNQ/M channels in dorsal root ganglion neurons. Further supporting this finding, another transcriptomic study using RNA from ASD mouse models identified *S1PR1* as a potential ASD-related target, reporting significantly different levels of expression and then further validated the results in post-mortem samples of autistic individuals^73^.

The two genomic blocks associated with ASD and linked to Native American ancestry in the main Multi-Ethnic approach are located near to each other on chr11q13.4 (0.59 Mb apart), encompassing several genes of interest. These include *SHANK2* and *DHCR7*, both previously implicated in ASD^74–76^, as well as potentially novel targets such as the *FOLR*^77^ gene family and *PHOX2A*^78^.

Notably, members of the *FOLR* family have been associated with Cerebral Folate Deficiency Syndrome (CFDS), an autosomal recessive neurodevelopmental condition characterized by reduced folate concentrations in the cerebrospinal fluid despite normal systemic folate levels. Variants in *FOLR1*, including loss-of-function variants^79,80^, missense variants^81^ and homozygous or compound heterozygous variants^82,83^ have been linked to CFDS. Public health measures such as mandatory folic acid fortification have substantially reduced folate deficiency worldwide; countries like the United States, Canada, and Australia report prevalence as low as 1.7%, compared with up to 23.8% in countries without fortification^84^. This locus has also been implicated in a previous GWAS examining heart rate recovery following exercise (10 seconds post-exercise)^85^. Moreover, some individuals with ASD have been reported to exhibit a significant heart rate decrease during physical testing compared to controls, suggesting a potential link between this locus and autonomic nervous system differences in ASD^86^. It is also noteworthy that the *FOLR1* gene encodes the alpha form of the folate receptor and it has been reported that when the receptor is compromised by auto-antibodies (or mutation) in individuals with ASD, treatment with d,l-leucovorin (folinic acid) may be beneficial^87^.

The AM block on chr11 encompassing the *SHANK2* and *DHCR7* genes remains particularly compelling, supported by strong statistical evidence from fine-mapping analyses (p=6.78×10⁻⁶). The region highlighted by fine-mapping lies between *SHANK2* and *DHCR7* and contains multiple distal enhancer-like elements identified by ENCODE, as well as CCCTC-binding factor (CTCF) binding sites. Both types of regulatory elements show evidence of chromatin interactions with these genes, suggesting a shared regulatory domain. Although this region is intragenic, the presence of histone modification peaks further supports its potential regulatory function (Figure 3). The LocusZoom^88^ plot also revealed GWAS Catalog hits within this region, specifically associated with serum 25-hydroxyvitamin D^89^ and vitamin D levels^90^. Although vitamin D-related traits are not specific to ASD, multiple studies have reported a potential biological link between vitamin D metabolism and ASD susceptibility ^91–93^.

This locus is further reinforced by the diverse ancestral distribution of CNV deletions identified in ASD-affected individuals, including seven individuals carrying the same recurrent CNV deletion (chr11:71,351,712–71,356,380). This recurrent CNV was detected in individuals of diverse ancestry, specifically, two of admixed ancestry, two of African ancestry, and three whose ancestry was not classified within the five groups considered in this study (Figure 3). According to gnomAD, this CNV is absent in most populations but present at low frequencies in Africans (2.01×10⁻³), and Admixed Americans (7.94×10⁻⁵). These findings underscore the critical importance of conducting association studies in diverse populations, as this variant would likely have been missed in European-only cohorts.

The refined analysis on Latin individuals also yielded significant results, identifying 16 loci that clustered into seven genomic blocks (Figure 4, Table 2, Supplementary Figure 3, Supplementary Table 2). These included genes previously implicated in ASD, such as *AGBL4* (SFARI score 2, no EAGLE score), and novel candidate genes, including *ARNTL* and *FAR1* (Table 2). *AGBL4* has been previously associated with ASD through the identification of multiple variants in unrelated families including compound heterozygous variants (compound heterozygous variants in this case are missense and the next missense variant is *de novo*)^94^, missense SNVs^95^ and CNVs^96^. These associations are functionally plausible given *AGBL4*’s role in central nervous system neuron development.

*ARNTL* also emerges as a strong ASD gene candidate due to its function as a circadian clock transcription factor involved in several ASD-related regulatory pathways. It upregulates the SFARI gene *PER2*^97^ (SFARI score 2, EAGLE score 6.75) and is repressed by another SFARI gene, *NR1D1*^98^ (SFARI score 2). Additionally, *ARNTL* is regulated by *RAI1* (SFARI score 1S, EAGLE score 12.25), highlighting a broader transcriptional network linked to circadian rhythm and neurodevelopment. Mouse model studies also support relevance, where *Arntl* haploinsufficiency leads to brain-wide mTOR hyperactivation and ASD-like behaviors^97^. Similarly, although *FAR1* is not currently listed as a SFARI gene, variants in this gene have been associated with a range of neurodevelopmental symptoms, including speech delay, intellectual disability, cataracts, growth retardation, and seizure disorders, typically without rhizomelia or skeletal abnormalities^99,100^.

In summary, we show that incorporating data from multiple ancestries through statistical methods such as AM can reveal new genetic associations and validate the importance of other findings.

## Data Availability

All data produced in the present work are contained in the manuscript

https://research.mss.ng/

https://base.sfari.org

https://www.internationalgenome.org/

https://www.cgen.ca/hostseq-databank-access-request

## Acknowledgments

We thank the families participating in MSSNG, SSC, and SPARK, as well as the resources provided by Autism Speaks and The Centre for Applied Genomics. We also acknowledge the Peruvian Genome Project (PGP) team for generating the dataset used exclusively as a reference panel for ancestry inference in this study. M.M.A was supported by the SickKids Restracomp Fellowship. S.W.S holds the Northbridge Chair in Pediatric Research at The Hospital for Sick Children and the University of Toronto. E.T.S is supported by the brazilian Conselho Nacional de Desenvolvimento Científico e Tecnológico (CNPq) and Fundação de Amparo à Pesquisa do Estado de Minas Gerais (FAPEMIG).

