## Supplementary Material for "Characterizing Features of the Genetic Architecture Underlying Autism from a multi-ancestry Perspective"

1. The Centre for Applied Genomics, The Hospital for Sick Children, Toronto, ON, Canada.
2. Genetics and Genome Biology Program, The Hospital for Sick Children, Toronto, ON, Canada.
3. Molecular Medicine Program, The Hospital for Sick Children, Toronto, ON, Canada
4. Núcleo de Pesquisas em Ciências Biológicas (NUPEB) Universidade Federal de Ouro Preto (UFOP)
5. Departamento de Genética, Ecologia e Evolução, Instituto de Ciências Biológicas, Universidade Federal de Minas Gerais, Belo Horizonte, MG, 31270-901, Brazil.
6. Children's Hospital Los Angeles. 4650 Sunset Blvd, Los Angeles, CA 90027
7. Institute for Genome Sciences - University of Maryland, Baltimore.
8. Institute for Health Computing - University of Maryland.
9. Department of Molecular Genetics and McLaughlin Centre, University of Toronto, Toronto, ON, Canada.

### Supplemental Figures and legends

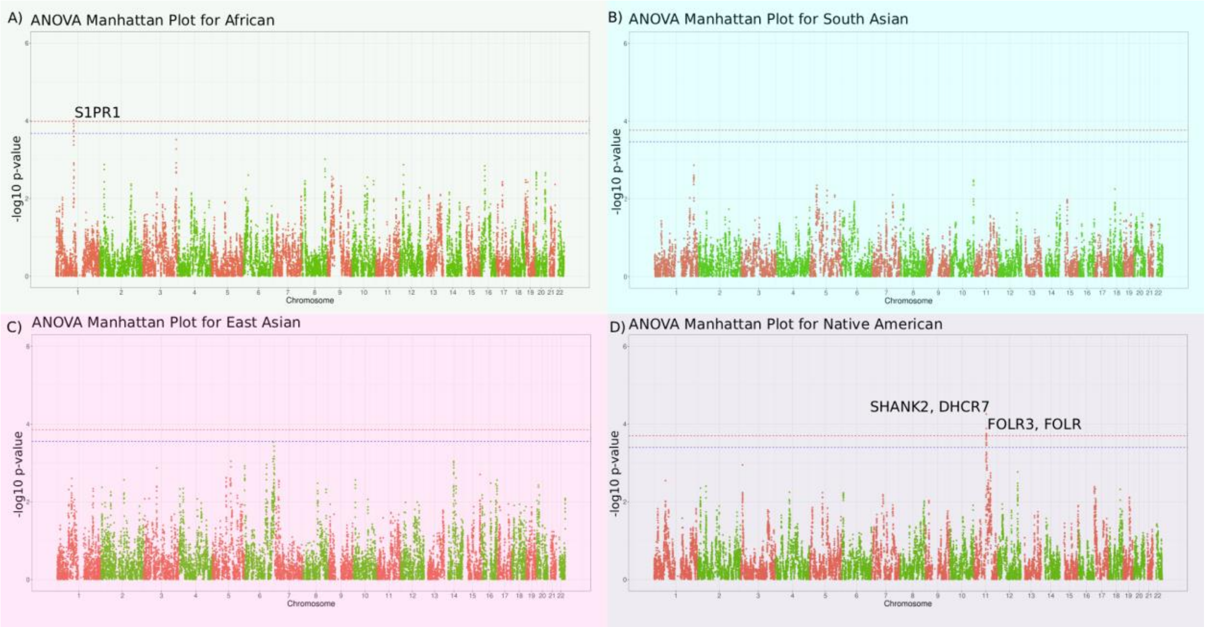


**Figure S1. Results for Multi-Ethnic ASD Admixture Mapping.** Manhattan plots of admixture mapping results for: A) African, B) South Asian ancestry, C) East Asian, and D) Native American tracts. The red horizontal line indicates the Bonferroni-corrected significance threshold (0.5/N effective tests), and the blue line represents the suggestive significance threshold (0.1/N effective tests).

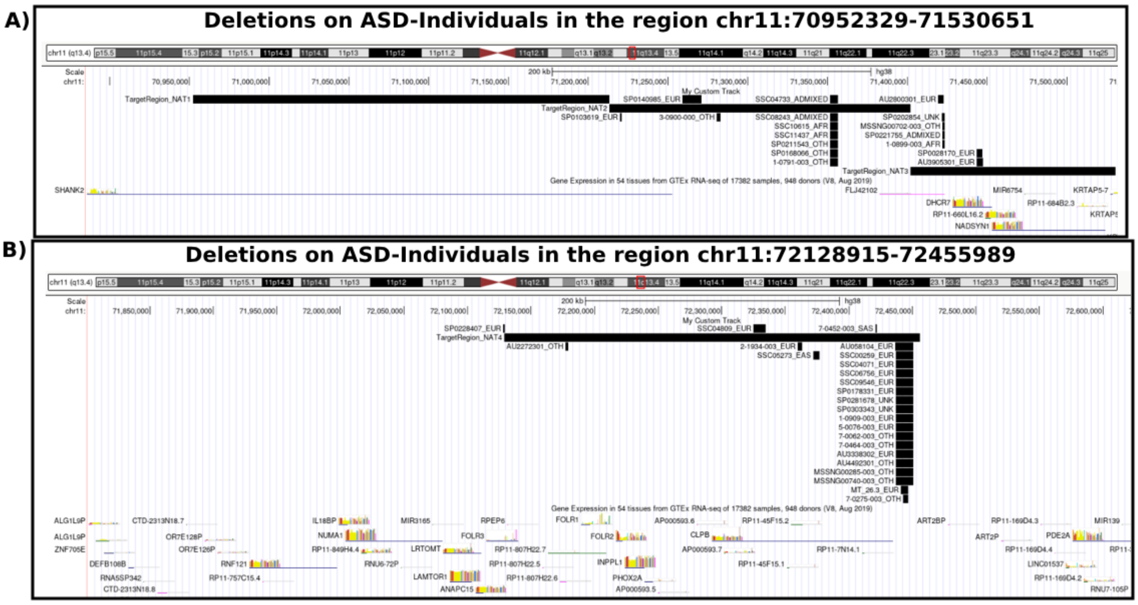


**Figure S2. CNV Deletions Overlapping Admixture Mapping Significant Loci Using Five Ancestry Sources.** A) Copy number deletions in ASD individuals overlapping the region chr11:70,952,329-71,530,651. B) Copy number deletions in ASD individuals overlapping the region chr11:72,128,914-72,455,989.

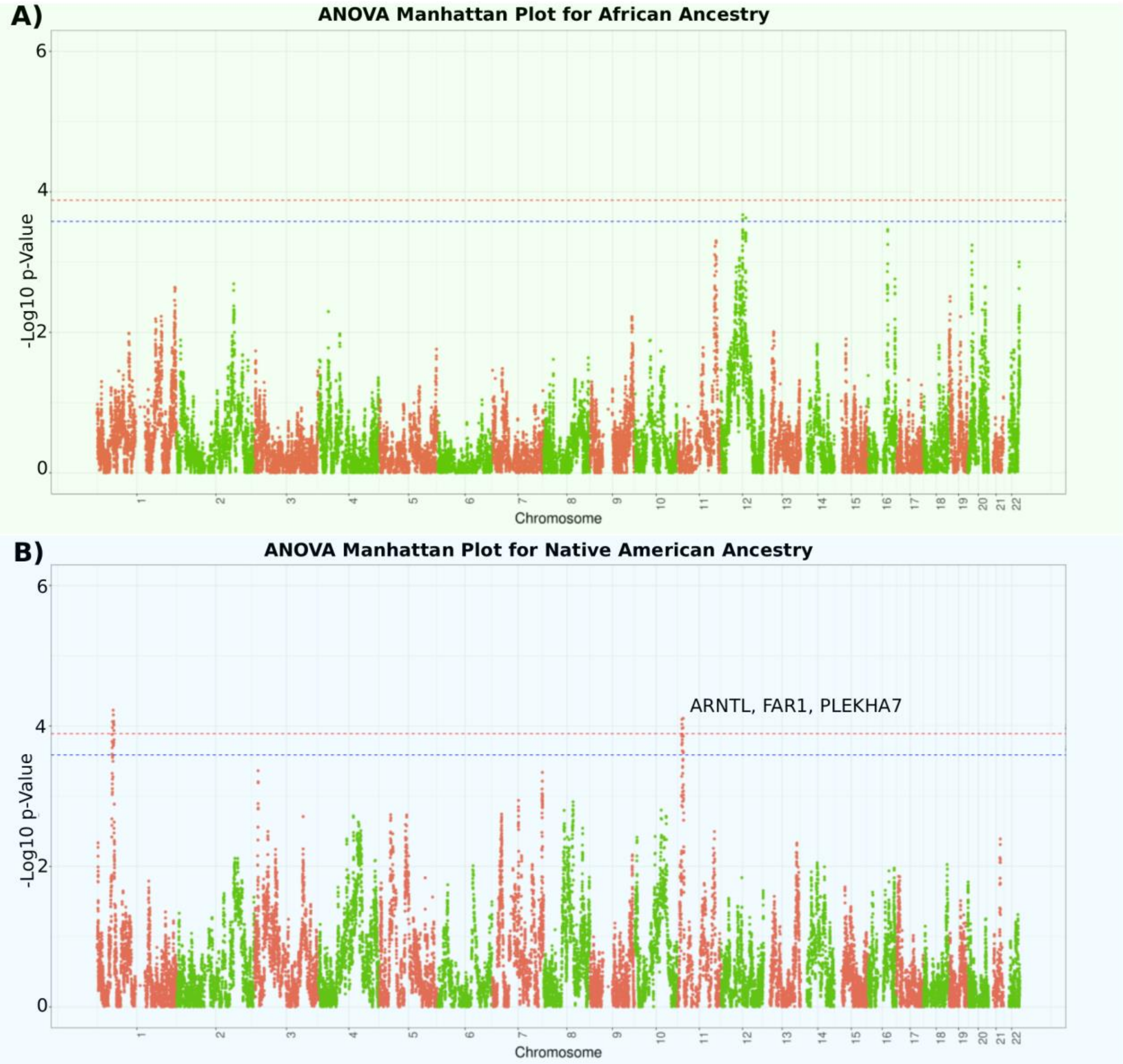


**Figure S3. Results for Refined Analysis in Latin American Populations.** Manhattan plots of admixture mapping results for: (A) African, (B) Native American tracts. The red horizontal line indicates the Bonferroni-corrected significance threshold (0.5/N effective tests), and the blue line represents the suggestive significance threshold (0.1/N effective tests).

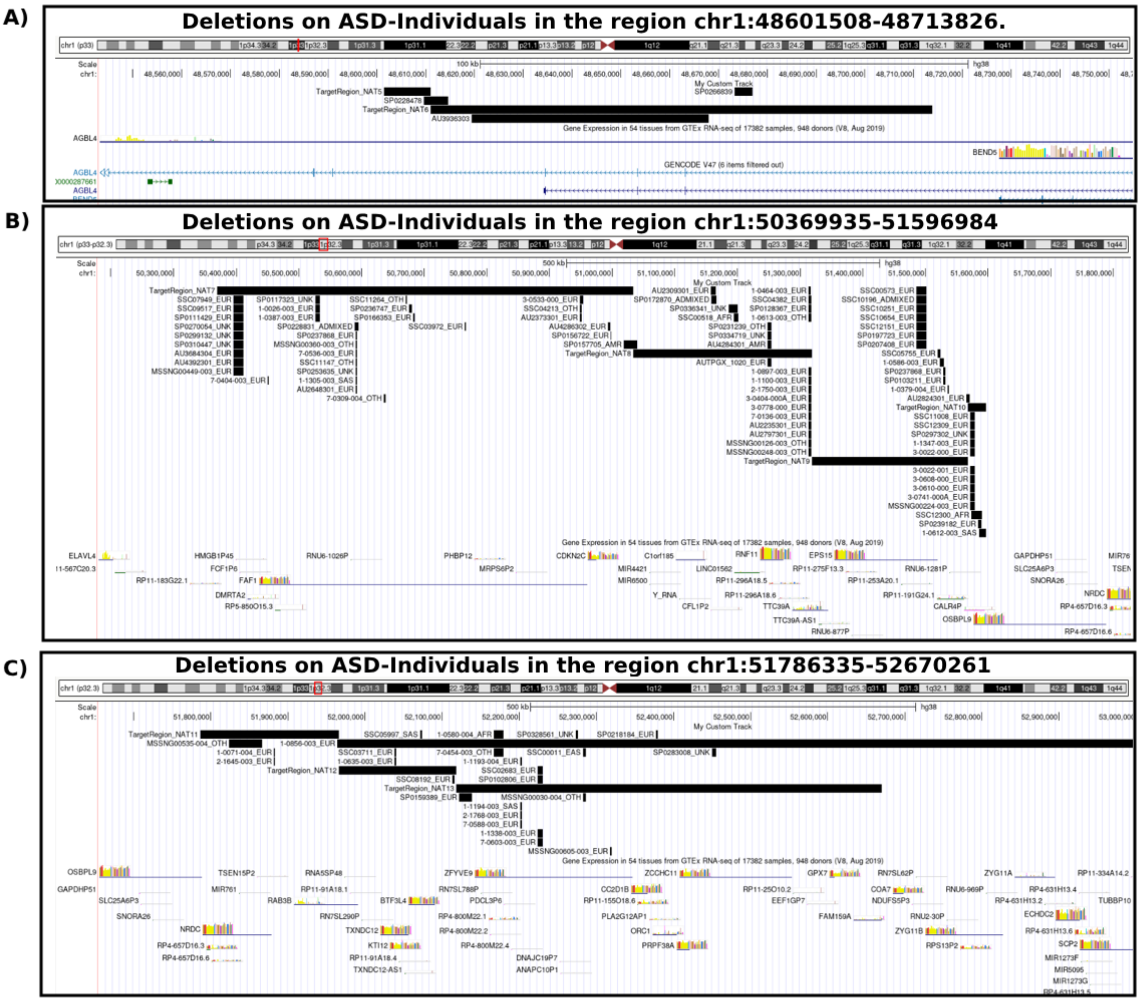


**Figure S4. CNV Deletions Overlapping Refined Analysis in Latin American Populations.** A) Copy number deletions in ASD individuals overlapping the region chr1:48,601,507-48,713,826. B) Copy number deletions in ASD individuals overlapping the region chr1:50,369,934–51,596,984. C)Copy number deletions in ASD individuals overlapping the region chr1:51,786,334–52,670,261.

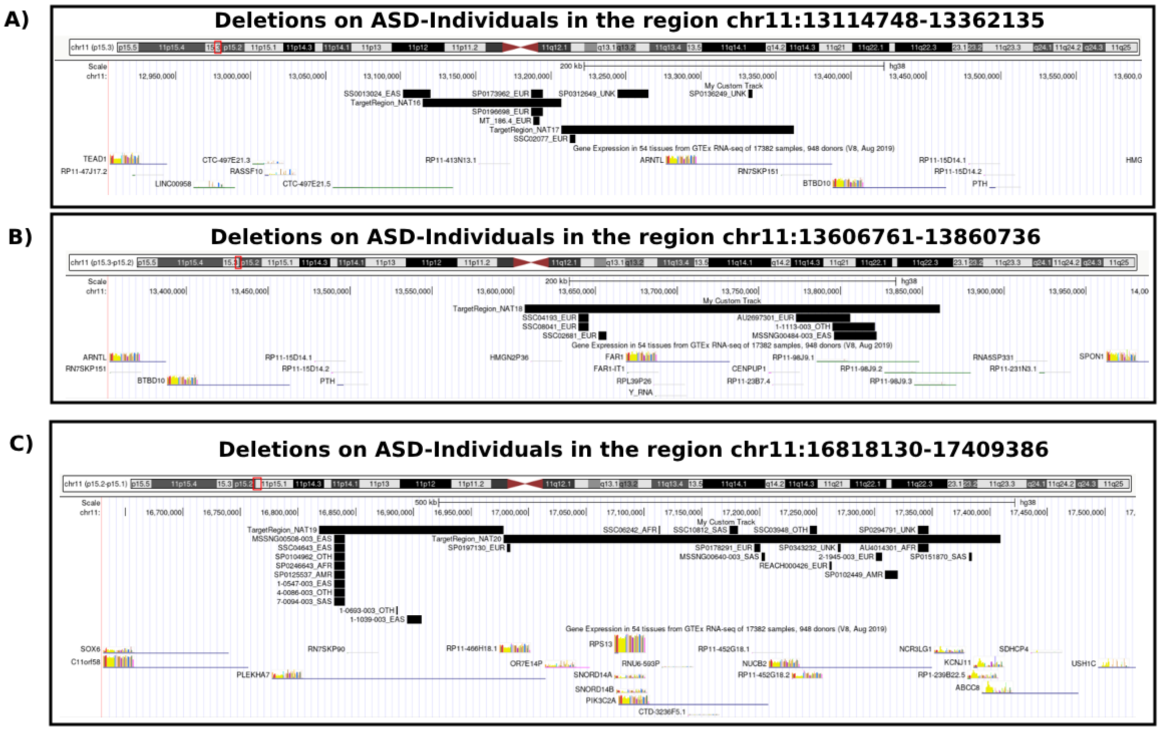


**Figure S5. CNV Deletions Overlapping Refined Analysis in Latin American Populations.** A) Copy number deletions in ASD individuals overlapping the region chr11:13,114,747-13,362,135. B) Copy number deletions in ASD individuals overlapping the region chr11:13,606,761-13,860,736. C)Copy number deletions in ASD individuals overlapping the region chr11:16,818,130-17,409,386.

### Supplemental tables

**Table S1. Prevalence of ASD across ethnic and ancestry groups.** This table summarizes the most recent ASD prevalence estimates stratified by ethnic or ancestry-related population groups. Reported values reflect population-level prevalence estimates as described in Salari et al. (2022)[1], and Shaw et al. (2025)[2]. Ethnic and geographical categories follow the terminology used in the original publications.

| Population tag | Brief population description | ASD prevalence (%) | Reference |
|---|---|---|---|
| Americas | Population-based studies from North, Central, and South America | 1.00 | Salari et al., 2022 |
| Europe | Predominantly European-ancestry populations from European countries | 0.50 | Salari et al., 2022 |
| Australia | Australian population cohorts | 1.70 | Salari et al., 2022 |
| Asia | East and South Asian populations | 0.40 | Salari et al., 2022 |

| | | | |
|---|---|---|---|
| Africa | African populations from population-based studies | 1.00 | Salari et al., 2022 |
| Hispanic or Latino | Individuals of Latin American ancestry, regardless of race | 3.30 | Shaw et al., 2025 |
| Non-Hispanic White | White individuals without Hispanic/Latino ethnicity | 2.77 | Shaw et al., 2025 |
| Asian or Pacific Islander | East Asian, Southeast Asian, or Pacific Island ancestry | 3.82 | Shaw et al., 2025 |
| Non-Hispanic Black or African American | Black individuals without Hispanic/Latino ethnicity | 3.66 | Shaw et al., 2025 |
| American Indian or Alaska Native | Indigenous peoples of North America | 3.75 | Shaw et al., 2025 |
| Multiracial | Individuals reporting more than one race/ancestry | 3.19 | Shaw et al., 2025 |
| All | Combined population estimate | 3.22 | Shaw et al., 2025 |

**Table S2. Summary of Admixture Mapping and Fine Mapping Results.** This table presents a summary of the admixture mapping results alongside findings from the refined analysis conducted in Latin American populations. It also includes results from the WGS-based Fine Mapping and the additional Fine Mapping incorporating the imputed EPIGEN dataset. Statistically significant p-values are highlighted in red. SNPs highlighted in green represent lead variants identified in both fine mapping analyses.

**Table S3. Summary Statistics from Admixture Mapping on Chromosome 1.** This table reports the p values for all loci tested on chromosome 1 across 5 ancestry sources: African, Native American, South Asian, East Asian, and European.

**Table S4. Summary Statistics from Admixture Mapping on Chromosome 11.** This table reports the p values for all loci tested on chromosome 11 across 5 ancestry sources: African, Native American, South Asian, East Asian, and European.

**Table S5. Summary Statistics Refined Analysis of Latin American Populations on Chromosome 1.** This table reports the p values for all loci tested on chromosome 1 across 3 ancestry sources: African, Native American, and European.

**Table S6. Summary Statistics Refined Analysis of Latin American Populations on Chromosome 11.** This table reports the p values for all loci tested on chromosome 11 across 3 ancestry sources: African, Native American, and European.

**Table S7. List of Mapped Genes from Admixture Mapping Results.** This table contains the list of genes mapped using the UCSC Table Browser, based on the genomic coordinates identified through admixture mapping.

*Datasets*

We used data from the Autism Speaks MSSNG Project[3,4], the Simons Simplex Collection (SSC)[5], and the Simons Foundation Powering Autism Research (SPARK) cohort[6] which collectively have genetic information for thousands of families with one or more individuals with ASD. From MSSNG, we used WGS data for 9,448 individuals mainly from North America including diverse ethnic backgrounds, after selecting only the variants that were shared with the reference data, all of them based on high-quality short-read sequencing (>30x coverage). These were 4,373 affected samples and 5,075 unaffected individuals. DNA quality assessment, quantitation, library preparation, and sequencing were performed as described in Yuen *et al.*, 2017[4], while read alignment and variant detection were performed as described in Trost *et al.*, 2022[7]. We also used WGS data from 2,419 affected individuals and 6,786 unaffected family members from SSC (https://base.sfari.org), who were recruited mostly from the United States of America. From SPARK, we had WGS data for 12,519 individuals from ASD families, with a total of 3,107 ASD probands. For controls, we used 10,000 samples from the cross-Canada Host Genome Sequencing Initiative (HostSeq)[8]. To refine the Native American–related findings, we also included 6,487 control samples from the Brazilian EPIGEN initiative[9], which includes samples from three Brazilian cohorts. We merged these datasets with autistic individuals and controls from North American cohorts (MSSNG, SSC, SPARK, and HostSeq) after filtering for individuals with self-reported or genetically inferred ethnicity (self-reported ethnicity was not always available) labeled as "American," "Admixed," "Hispanic," "Other," or "Unknown." We included individuals labeled as "Other" or "Unknown" in this initial filter, to avoid excluding participants with potential Native American or admixed ancestry, which may be underreported or misclassified in existing records.

As reference data, we used samples from the 1000 Genomes Project phase 3 data (1KGP)[10,11], including 404 individuals of European ancestry (GBR=91N, IBS=107, TSI=107N, CEU=99N), 425 with African ancestry (YRI=108N, ASW=56N, MSL=78N, ESN=95, LWK=88N), 405 with East Asian ancestry (CHS=107N, CHB=103N, CDX=91N, JPT=104N), 402 South Asian ancestry (ITU=102N, GIH=102N, PJL=96N, STU=102N) and 64 individuals from Mexico with Native American ancestry (MXL). We also used as reference, 358 Peruvians from the Peruvian Genome Project (PGP)[12,13], with Native American Ancestry.

*Quality Control*

In addition to the existing quality control procedures applied to the MSSNG, SSC, HostSeq, EPIGEN, 1KGP, and PGDP datasets, we implemented an additional set of filters specific to our admixture mapping analysis (Figure 1). We began by selecting SNPs present in the reference panel (1KGP and PGP) and retained only biallelic variants (bcftools view --max-alleles 2). This step was necessary because widely used tools for ancestry inference and association testing, such as PLINK, are designed to handle only biallelic markers. Separately for each dataset, we applied a standardized QC pipeline[14], which included the removal of SNPs with missing data, strand-ambiguous alleles, and the annotation of variants using dbSNP151[15]. We then merged the affected samples from the ASD datasets (MSSNG=4,373; SSC=2,419; SPARK=3,107) with the controls and reference data, resulting in a total of 1,533,979 variants and 21,980 samples. After inferring the kinship matrix using KING[16] , implemented via PLINK2 (--make-king-table), we applied a second-degree relatedness filter using a network-based pruning method implemented in the NAToRA software[17] resulting in a final dataset of 20,794 samples with 1,533,979 variants.

*Population Structure*

For population structure analysis, we used the dataset after applying additional variant-level filters, including Hardy-Weinberg equilibrium (HWE $p > 1e-6$), minor allele frequency ($MAF > 0.05$), exclusion of regions under natural selection, such as the human leukocyte antigen (HLA) region (hg38:chr6:27032221-35032223, chr2:134242429-136242430, chr8:6142478-16142491, chr17:41843748-46922634, chr2:108383544-109383544, and chr7_KI270803v1_alt:339348-517347)[18–20], and linkage disequilibrium pruning ($r^2 < 0.1$). After these filters, 228,904 variants remained. To estimate global ancestry, we ran ADMIXTURE[21] with 10 replicates and K = 5, corresponding to five continental reference populations: Europeans, Africans, East Asians, South Asians, and Native Americans (Figure 2-A,B). We also performed principal component analysis (PCA) using SNPRelate[22].

*Local Ancestry Inference*

We phased our genome-wide genotype data (1,533,979 variants and 21,980 samples) with SHAPEIT4[23] using the GRCh38 genetic map[24] and the Markov Chain Monte Carlo (MCMC) parameters for 10 burn-in iterations, followed by four paired runs of pruning and burn-in, and finally 10 main iterations of sampling. Local ancestry inference was performed using RFMix v2[25]. EM (Expectation-Maximization) iterations were enabled by default and used to refine ancestry inference across the genome. For the subsequent analysis, we used the RFMix.msp.tsv output, which contains the most likely assignment ancestry per conditional random field (CRF) and the marginal probabilities of each reference population being the ancestral source of the corresponding CRF point.

*Sample Selection*

To prioritize admixed populations, we chose samples from both autistic individuals and control datasets, ensuring that none exceeded an 80% proportion in any ancestry source (Figure 2). Additionally, for the subsequent analysis, we selected samples from the reference populations (1KGP, and PGDP) with more than 80% representation from a single ancestry source to ensure their suitability. After this filter we had 1,747 autistic individuals; 1,606 controls and 2,043 reference samples (Figure 2-D).

We then applied a case/control matching algorithm, selecting the closest control sample for each case using an adapted version of PCAmatchR[26] (Figure 1-A and Figure2-D,E). All controls and reference samples were split into 32 subgroups. For each subgroup, pairwise weighted Mahalanobis distance was calculated between controls and reference samples with all autistic individuals using the first 5 principal components. The top 20 autistic individuals with the closest weighted Mahalanobis distances were selected for each control and reference sample. Matched results greater than the mean Mahalanobis distance plus 1 standard deviation (1SD) were removed. All matched pairs were selected to retain for the case versus control analysis. If two autistic individuals were optimally matched to the same control, the case with the smaller weighted Mahalanobis distance was kept, and the other removed to ensure one-to-one case/control matching. After this filter we had our final datasets for the admixture mapping analysis that had 1,033 autistic individuals; 1,033 controls and 2,043 reference samples (Figure 2-D).

*Admixture mapping*

For each segment inferred from RFMix, we assessed the number of African, European, East Asian, South Asian, and Native American tracts (0, 1, or 2). We then used logistic regression to test the association between ASD and the number of each ancestry tracts, with ASD as a binary trait. To avoid the dummy variable trap and ensure model identifiability, we set European ancestry as the reference category and excluded it from the regression model.

The logistic regression models were adjusted for sex and the global ancestry proportions as fixed-effect covariates[27,28]. Model comparisons were performed using likelihood ratio tests (ANOVA, Chi-squared test) to determine whether adding local ancestry at each locus significantly improved model fit.

The full logistic regression model can be expressed mathematically as:

$$log\left(\frac{P(Y=1)}{1-P(Y=1)}\right) = \beta_0 + \sum_{j=1}^{2} \beta_j G_j + \sum_{k=1}^{5} \gamma_k A_k + \delta \cdot SEX$$

Where:

- $Y_i$ = ASD status for individual $i$
- $G^{(j)}$ = local ancestry dosage from ancestry $j$ at the tested tract (excluding the baseline)
- $A_k$ = global ancestry proportions for the 5 inferred components
- $SEX_i$ = sex of individual $i$
- $\beta_0$ = intercept
- $\beta_j$, $\gamma_k$, $\delta$ = regression coefficients

*Significance Threshold*

Since our association tests are based on tracts in the genome rather than individual SNPs, the conventional genome-wide significance threshold $5\times10^{-8}$ is not applicable. To determine an appropriate significance threshold for our admixture mapping analysis, we applied a Bonferroni correction by dividing 0.05 by the number of effective tests.

The number of effective tests was estimated by calculating a correction factor that accounts for the correlation structure across the genome. Specifically, we started with the total number of ancestry tracts (i.e., the raw or unadjusted number of tests prior to accounting for correlation), scaled this by the sample variance, and then divided by the estimated spectral density at frequency zero. The spectral density was estimated using the CODA package[29], which applies an autoregressive (AR) model via the “ar function”. This estimate is derived from the prediction variance (variance unexplained by the model) and the coefficients of the AR model. Additional details can be found in the CODA package documentation[30]. The number of effective tests (Neff) based on the ancestry tracts found is as follows: African=476.78, Native American=251.04, South Asian=293.58, and East Asian=355.29. Dividing 0.05 by Neff yields the following Bonferroni-corrected significance thresholds: African=$1.05\text{x}10^{-4}$, Native American=$1.99\text{x}10^{-4}$, South Asian=$1.70\text{x}10^{-4}$ and East Asian=$1.41\text{x}10^{-4}$.

*Fine Mapping*

For fine mapping analyses, we utilized all unrelated WGS samples, regardless of ancestry, resulting in a combined dataset of 18,641 individuals (9,083 autistic individuals and 9,558 controls). Variants located within 250,000 base pairs upstream and downstream of the target regions identified in the admixture mapping results were extracted. We performed variant-level normalization and quality control (QC) on genomic data from four WGS cohorts: MSSNG, SSC, SPARK, and HostSeq, focusing on chromosomes 1 and 11. Variant call format (VCF) files were normalized against the GRCh38 reference genome using bcftools norm, and only biallelic variants were retained using bcftools view[31]. Variants were filtered to retain those with a QUAL score >30 and a minor allele frequency (MAF) >0.05. Annotations were updated with bcftools +fill-tags, and only variants passing internal quality filters (PASS) were kept.

After variant-level filtering, individuals and variants with more than 5% missing data were removed using PLINK2[32]. dbSNP identifiers were updated using dbSNP151[15]. Additional QC steps included testing for Hardy-Weinberg equilibrium (HWE; p<1e-5, mid-p adjustment) and differential missingness between cases and controls (--test-missing in PLINK[33]). Variants failing either filter were excluded, and final cleaned binary genotype files were generated using PLINK2. The cleaned datasets from all cohorts were merged, resulting in 5,731 variants on chromosome 1 and 8,299 on chromosome 11.

Association testing was performed using logistic regression in PLINK, adjusting for sex and the first five principal components (PCs) to account for population structure. Covariates were variance-standardized (--covar-variance-standardize) to place them on comparable scales, improving model stability and interpretability. Confidence intervals were calculated at 95% (--ci 0.95).

*Locus interpretation and gene prioritization*

To interpret potential ASD-associated loci and prioritize candidate genes, we annotated the genomic regions of interest using the UCSC Table Browser tool[34]. We examined fine-mapping results using LocusZoom[35] to visualize regional association signals. To assess population-level allele frequencies of lead SNPs identified through fine-mapping, we incorporated reference data from the 1000 Genomes Project[11].

13. Borda, V. *et al.* The genetic structure and adaptation of Andean highlanders and Amazonians are influenced by the interplay between geography and culture. *Proc Natl Acad Sci U S A* **117**, 32557–32565 (2020).
14. GitHub - ldgh/MosaiQC-public: SmartCleaning (now called MosaiQC) is a script developed by Thiago Peixoto Leal to clean and perform QC of genotyping array data using PLINK. It automates the different steps. If interested in using SmartCleaning, contact the LDGH/Mosaico Translational Genomics team. *GitHub* https://github.com/ldgh/MosaiQC-public.
15. Sherry, S. T. *et al.* dbSNP: the NCBI database of genetic variation. *Nucleic Acids Res* **29**, 308–311 (2001).
16. Manichaikul, A. *et al.* Robust relationship inference in genome-wide association studies. *Bioinformatics* **26**, 2867 (2010).
17. NAToRA, a relatedness-pruning method to minimize the loss of dataset size in genetic and omics analyses. *Comput. Struct. Biotechnol. J.* **20**, 1821–1828 (2022).
18. Price, A. L. *et al.* Long-range LD can confound genome scans in admixed populations. *Am J Hum Genet* **83**, 132–5; author reply 135–9 (2008).
19. Turner, S. *et al.* Quality Control Procedures for Genome Wide Association Studies. *Current protocols in human genetics / editorial board, Jonathan L. Haines ... [et al.]* **CHAPTER**, Unit1.19 (2011).
20. Anderson, C. A. *et al.* Data quality control in genetic case-control association studies. *Nature Protocols* **5**, 1564–1573 (2010).
21. Alexander, D. H., Novembre, J. & Lange, K. Fast model-based estimation of ancestry in unrelated individuals. *Genome Res.* **19**, 1655–1664 (2009).
22. Zheng, X. *et al.* A high-performance computing toolset for relatedness and principal component analysis of SNP data. *Bioinformatics* **28**, 3326–3328 (2012).
23. Delaneau, O., Zagury, J.-F., Robinson, M. R., Marchini, J. L. & Dermitzakis, E. T. Accurate, scalable and integrative haplotype estimation. *Nat. Commun.* **10**, 1–10 (2019).

24. shapeit4/maps at master · odelaneau/shapeit4. *GitHub* https://github.com/odelaneau/shapeit4/tree/master/maps.
25. Maples, B. K., Gravel, S., Kenny, E. E. & Bustamante, C. D. RFMix: a discriminative modeling approach for rapid and robust local-ancestry inference. *Am. J. Hum. Genet.* **93**, 278–288 (2013).
26. Brown, D. W., Myers, T. A. & Machiela, M. J. PCAmatchR: a flexible R package for optimal case-control matching using weighted principal components. *Bioinformatics* **37**, 1178–1181 (2021).
27. Reynolds, K. M. *et al.* Ancestry-driven metabolite variation provides insights into disease states in admixed populations. *Genome Medicine* **15**, 1–13 (2023).
28. Horimoto, A. R. V. R., Xue, D., Thornton, T. A. & Blue, E. E. Admixture mapping reveals the association between Native American ancestry at 3q13.11 and reduced risk of Alzheimer's disease in Caribbean Hispanics. *Alzheimer's Research & Therapy* **13**, 1–14 (2021).
29. Plummer, M., Best, N., Cowles, K. & Vines, K. CODA: convergence diagnosis and output analysis for MCMC. *R News* **6**, 7–11 (2006).
30. effectiveSize function - RDocumentation. https://www.rdocumentation.org/packages/coda/versions/0.19-4/topics/effectiveSize.
31. Danecek, P. *et al.* Twelve years of SAMtools and BCFtools. *Gigascience* **10**, (2021).
32. Chang, C. C. *et al.* Second-generation PLINK: rising to the challenge of larger and richer datasets. *GigaScience* **4**, 7 (2015).
33. Purcell, S. *et al.* PLINK: a tool set for whole-genome association and population-based linkage analyses. *Am J Hum Genet* **81**, 559–575 (2007).
34. UCSC Genome Browser Home. https://genome.ucsc.edu/.
35. Boughton, A. P. *et al.* LocusZoom.js: interactive and embeddable visualization of genetic association study results. *Bioinformatics* **37**, 3017–3018 (2021).
